# Sex Differences in the Role of Sleep on Cognition in Older Adults

**DOI:** 10.1101/2024.01.08.24300996

**Authors:** Yumiko Wiranto, Catherine Siengsukon, Diego R. Mazzotti, Jeffrey M. Burns, Amber Watts

## Abstract

**Study Objectives:** The study aimed to investigate sex differences in the relationship between sleep quality (self-report and objective) and cognitive function across three domains (executive function, verbal memory, and attention) in older adults.

**Methods:** We analyzed cross-sectional data from 207 participants with normal cognition or mild cognitive impairment (89 males and 118 females) aged over 60. The relationship between sleep quality and cognitive performance was estimated using generalized additive models. Objective sleep was measured with the GT9X Link Actigraph, and self-reported sleep was measured with the Pittsburgh Sleep Quality Index.

**Results:** We found that females exhibited stable performance of executive function with up to about 400 minutes of total sleep time, with significant declines in performance (*p* = 0.02) when total sleep time was longer. Additionally, a longer total sleep time contributed to lower verbal memory in a slightly non-linear manner (*p* = 0.03). Higher self-reported sleep complaints were associated with poorer executive function in females with normal cognition (*p* = 0.02). In males, a positive linear relationship emerged between sleep efficiency and executive function (*p* = 0.04), while self-reported sleep was not associated with cognitive performance in males with normal cognition.

**Conclusions:** Our findings suggest that the relationships between sleep quality and cognition differ between older males and females, with executive function being the most influenced by objective and self-reported sleep. Interventions targeting sleep quality to mitigate cognitive decline in older adults may need to be tailored according to sex, with distinct approaches for males and females.

**Statement of Significance:** This research is significant as it addresses the gap in a previously underexplored area of sex differences in the interplay between sleep quality and cognitive function in older populations. By examining both objective and self-reported sleep measures across three cognitive domains, the study provides valuable insights into how sleep impacts various cognitive domains differently in older males and females. Notably, the findings suggest that interventions to improve sleep quality and potentially mitigate cognitive decline may need to be sex-specific.

## Introduction

The impact of aging on sleep and cognition is well documented in the literature. Aging has been associated with fragmented sleep and decreased sleep quality, which could exacerbate cognitive decline that occurs with aging ^1^. However, the literature on sex differences in sleep and its interaction with cognition in older adults is still limited. Females experience dramatic hormonal and physiologic developmental changes (puberty, pregnancy, and menopause) across the lifespan that contribute to alterations in sleep patterns and a higher number of reported sleep complaints ^2^. Moreover, females have twice the risk of developing Alzheimer’s disease (AD), for which poor sleep is a risk factor ^3,4^. Understanding how males and females differ in their sleep health and the implications of these differences on cognition is vital for designing effective interventions and prevention strategies for AD.

Advancing age is associated with physiological changes, among which are the alterations of sleep architecture (i.e., stages of sleep) and circadian rhythm. Older adults have a shorter slow-wave sleep duration and activity, as well as shorter rapid eye movement sleep and more frequent awakenings during the night ^5,6^. Additionally, they tend to go to bed earlier in the evening and wake up earlier in the morning ^7^. These changes contribute to the high prevalence of sleep disturbances experienced by this population. Previous studies reported that up to 75% of older adults experienced sleep difficulty ^8–10^. Because one of the roles of sleep is to clear out neurotoxic waste products produced during the day, poor sleep has been found to exacerbate cognitive decline and neuropathological markers of AD, such as beta-amyloid and tau deposition^11–13^.

The investigation of the interrelationships between aging, sleep, and cognition has been relatively inconclusive and marked by variability in findings, potentially due to methodological variations and differences in operationalizing sleep metrics across studies. In older adults with normal cognition, both short (< 6 hours) and long (> 9 hours) sleep durations have been linked to reduced global cognition and memory ^14,15^. However, Sabeti et al. ^16^ observed this association solely in older adults with cognitive impairment. In a two-year longitudinal study, it was found that older females who slept for only four hours exhibited a more significant cognitive decline compared to those who slept for seven hours. Notably, individuals who slept for nine hours did not experience any cognitive decline during the same period ^17^. Conversely, Faubel et al. ^18^ indicated that older adults who slept for 11 hours demonstrated lower cognitive performance compared to those who slept for seven hours. In addition to global cognition and memory, sleep disturbances have been reported to impact other cognitive domains, including attention and executive function, in older populations. Specifically, Blackwell et al. ^19^ and Wilckens et al. ^20^ revealed a negative correlation between frequent nighttime awakenings and performance on executive function and attention. However, Wilckens et al. ^20^ did not find a significant association between sleep duration and cognitive performance in their study.

Biological sex is another major factor that influences sleep. Research investigating sex differences in self-reported sleep quality suggests that middle-aged and older females take longer to fall asleep, have shorter sleep duration, and have lower sleep efficiency ^21,22^. However, this result is contradicted by studies looking at objective sleep quality between older males and females using actigraphy and polysomnography (PSG). A meta-analysis including studies on sleep characteristics across three countries reported that older females had longer sleep duration and higher sleep efficiency based on actigraphy data ^23^. Studies using PSG show that reduced slow-wave sleep due to age is more pronounced in males than females ^24,25^. The variation between self-reported and objective sleep quality may be attributed to sex-related differences in sleep perception and symptom reporting. Recent research suggests a stronger link between objective and self-reported sleep quality in females compared to males ^26,27^, which may reflect a closer alignment between females’ self-reported sleep experiences and objective sleep parameters. Additionally, it raises questions about whether the current criteria for defining good objective sleep quality apply equally to both sexes, suggesting a need for distinct criteria for females ^28,29^.

Regarding the cognitive implications of poor sleep, limited research has been conducted to investigate sex differences in this topic. Notably, it has been observed that sleep deprivation affects working memory to a greater extent in young females compared to males ^30,31^. However, whether the same findings apply to older populations is still not well understood. This lack of research leaves an important gap in our understanding of how sleep impacts cognitive function across the lifespan. Some possible mechanistic explanations for sex differences in this relationship include depletion in sex steroids (estradiol) due to menopause that could adversely affect cognition, specifically memory processing ^32^. Moreover, since sleep is known to influence memory performance, the connection between reduced estradiol levels due to menopause and memory might be further intensified by poor sleep.

In this study, we investigated sex differences in the relationship between sleep quality (self-reported and objective measures) and cognitive performance in a clinical cohort of older individuals. Because the research on this topic is limited, the goal of this paper was to generate hypotheses and identify potential patterns that can be explored further in subsequent studies. We were also interested in exploring the relationship between these two sleep quality measures and in assessing whether these relationships varied between males and females. Lastly, we explored whether cognitive performance was better explained by an objective measure, a self-reported measure, or a combination of both.

## Materials and Methods

### Participants

Participants were recruited from the University of Kansas Alzheimer’s Disease Research Center (KU-ADRC) Clinical Cohort, an ongoing longitudinal observational study to support regional and national research on aging, cognition, and AD. This study collects demographic, medical, psychological, and cognitive data annually, and imaging and biospecimen data as needed if participants opt in. Participants could opt to partake in the Physical Activity and Sleep Study (PASS), a sub-study of the Clinical Cohort initiated on July 29, 2015, in which participants completed self-report sleep questionnaires annually and wore an actigraphy every two years until May 27, 2021. During this time, the KU-ADRC Clinical Cohort enrolled over 1,000 participants. To be eligible for the Clinical Cohort study, participants had to be at least 60 years old, native English speakers, and not exhibit the following conditions: significant depressive symptoms, untreated thyroid dysfunction, visual or auditory impairment, active (< 2 years) ischemic heart disease, and uncontrolled insulin-dependent diabetes mellitus.

In the current analysis, we included 207 participants with normal cognition (NC) or mild cognitive impairment (MCI) as indicated by clinical dementia rating (CDR) scores of 0 and >0, respectively. Among these participants, 161 exhibited NC, while 46 had MCI with a CDR of 0.5 (*n* = 36) or 1 (*n* = 10). The research was approved by the KU-ADRC’s Institutional Review Board, and all participants provided written informed consent separately for both the KU-ADRC Clinical Cohort and the PASS study.

### Sleep Quality Measures

#### Actigraphy

ActiGraph GT9X Link accelerometers (Pensacola, FL) are body-worn monitors that measure objective sleep patterns. Our study used placement on the non-dominant wrist to estimate sleep parameters. The GT9X devices were programmed to collect data at a sample rate of 30 Hz. ActiLife software version 6.13.2 or 6.13.4 (ActiGraph, LLC) was used to process and analyze the raw actigraphy data using the Cole-Kripke algorithm ^33^. Wear time was measured using the Choi algorithm, and a valid wear time included a minimum of ten hours per day, at least one weekend day, and a minimum of four days ^34^. Participants were also provided with a sleep diary to document the timing and length of their sleep during the day and at night. They were asked to note how long it took them to fall asleep, record any instances of waking up during the night, and track the time they woke up and got out of bed.

We included sleep efficiency (SE) and total sleep time (TST) in this analysis due to the prevalent occurrence of sleep disturbances and variations in sleep duration and sleep efficiency among older adults. Wake after sleep onset was not included because it was highly correlated with SE (*r* = -0.95) in our sample. SE refers to the percentage of total time spent asleep during the total time in bed (Time asleep / Time in Bed). TST refers to the time spent asleep in bed (in minutes) at night. TST was calculated by subtracting the duration of falling asleep and awakening from the total amount of time in bed (in minutes; Out of Bed Time – In Bed Time). A higher TST means a longer sleep duration. For each night’s data, sleep intervals recorded by ActiGraph were compared with the participants’ sleep diaries to check for discrepancies. When the sleep diary entries were closely aligned (within a 30-minute margin) with the ActiGraph-identified sleep period, the sleep period identified by ActiGraph was used. In cases where the discrepancy exceeded 30 minutes, the researchers manually evaluated the sleep data in ActiLife by looking for significant changes in activity levels. This manual assessment was also automatically initiated whenever participants did not submit or complete their sleep diaries.

#### The Pittsburgh Sleep Quality Index

The Pittsburgh Sleep Quality Index (PSQI) is a self-report questionnaire consisting of 10 items measuring sleep quality over the past month. The total score of PSQI is calculated by summing its seven subscales, with scores ranging from 0 (indicating "good sleep quality") to 21 (indicating "poor sleep quality"). The PSQI is a reliable measure to distinguish between good and poor sleepers (PSQI > 5) with a sensitivity of 89.6% and a specificity of 86.5% ^35^. It is also valid to use for clinical and research purposes. In exploratory analyses, we also selected two of these subscales (scores range from 0 to 3), specifically sleep efficiency and duration, for comparison with SE and TST as measured by actigraphy.

### Cognitive Performance

As part of the Clinical Cohort study annual evaluation, participants completed a cognitive test battery, specifically the National Alzheimer’s Coordinating Center (NACC) Uniform Data Set (UDS), and several additional tests. The UDS was designed to measure cognitive performance in mild cognitive impairment or dementia due to AD by the Alzheimer’s Disease Centers Clinical Task Force established by the National Institute on Aging. The subtests included in the present analyses were the Wechsler Adult Intelligence Scales-Revised (WAIS-R; Digit Symbol Substitution Test) ^36^, two subtests from the Wechsler Memory Scales-Revised (WMS-R; Letter Number Sequencing and Digit Span Forward and Backward) ^37^, the Stroop Test (interference condition) ^38^, the Craft Story 21 Immediate and Delayed recall ^39^, the Free and Cued Selective Reminding Test ^40^, two tests of semantic verbal fluency (animal and vegetable naming) ^41^, and Trail Making Test B ^42^. The administration procedures of the subtests are described in their respective references.

The present study used cognitive factor scores derived from a confirmatory factor analysis. This approach reduces type 1 error resulting from multiple testing and improves measurement accuracy by combining common variance across multiple subtests while accounting for measurement error. The details of this analysis can be found elsewhere ^43^. The result yielded three factors: verbal memory (immediate and delayed logical memory, selective reminding test trials sum), attention (digits forward, digits backward, letter-number sequencing), and executive function (category fluency sum of animals and vegetables, Stroop color word interference, Trail Making Test B, and digit symbol substitution test).

### Statistical Analysis

We compared demographics (age and education) between males and females using an independent t-test. Analysis of covariance (ANCOVA) was used to compare the three cognitive domains and sleep variables (TST, SE, and PSQI) between sexes, adjusting for age and CDR, with education included as a covariate exclusively for the cognitive domains. We implemented Bonferroni correction to correct the Type I error rate to adjust for multiple comparisons at *p* < 0.008. We implemented generalized additive models (GAMs) with the mgcv package in R to analyze the relationship between sleep and each cognitive domain (executive function, verbal memory, and attention). Because previous studies have suggested that males and females have different sleep patterns and perceptions of sleep quality, we chose to perform the analyses separately for males and females to allow for clearer interpretations. We selected GAMs because previous research has shown that sleep and cognition were non-linearly correlated in older adults ^44^. GAMs are suitable for accounting for non-parametric and non-linear relationships. The spline fitting, a method to find the best fit of the data points, was estimated using Restricted Maximum Likelihood (REML). REML is particularly adept at reducing bias in the estimation of variance components. It adjusts for the degrees of freedom used by the fixed effects, allowing for more accurate and less biased estimates of the random effects variance. There was 14% (*n* = 29) of missing data in the PSQI variable which was imputed with multiple imputation chained equation with the mice package in R. All variables were included in the imputation, except for the dependent variables (i.e., the three cognitive domains). The imputation was performed by dividing the dataset into two: cognitively healthy and cognitively impaired groups. This was done because previous research has shown that cognitively impaired older adults have skewed awareness resulting in inaccurate self-reporting ^45^. Other variables, such as age, years of education, and CDR, that affect cognitive performance were added as covariates.

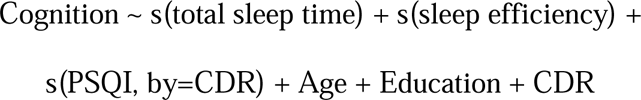

### Exploratory Analysis

We conducted Spearman’s correlation analysis to explore the relationship between sleep parameters (TST and SE) measured by actigraphy and PSQI, and to examine potential variations based on sex and CDR status. We also performed model comparisons to assess which model best explains the relationship between sleep and cognition. Specifically, we compared models based on actigraphy-measured sleep parameters, self-reported sleep parameters from the PSQI, a combination of both, and the original model. All models were estimated using GAM, and model comparisons were based on the Akaike Information Criterion (AIC).

<u>Model I:</u> Cognition ∼ s(total sleep time) + s(sleep efficiency) + Age + Education + CDR

<u>Model II:</u> Cognition ∼ PSQI sleep duration + PSQI sleep efficiency + s(PSQI) + Age + Education + CDR

<u>Model III</u>: Cognition ∼ s(total sleep time) + s(sleep efficiency) + PSQI sleep duration + PSQI sleep efficiency + s(PSQI) + Age + Education + CDR

<u>Model IV</u>: Cognition ∼ s(total sleep time) + s(sleep efficiency) + s(PSQI, by=CDR) + Age + Education + CDR

## Results

The summary of participants’ demographics is presented in Table 1. Most participants were White (93.70%), college-educated (16.4 years), and had normal cognition (i.e., CDR = 0; 77.78%). Our results revealed that females outperformed males in verbal memory (β = 0.32, *p* = 0.03) accounting for age, educational level, and CDR score. However, the sex differences in verbal memory were no longer significant after correcting for Type I errors using Bonferroni correction at *p* < 0.008. We did not find significant differences in executive function, attention, TST, SE, and PSQI between males and females.

**Table 1.** Descriptive Statistics on Demographics, Sleep, and Cognitive Performance.

| Participants characteristics | Males (*n* = 89) |  | Females (*n* = 118) |  | Total (*N*= 207) |  |
| --- | --- | --- | --- | --- | --- | --- |
|  | *M* | *SD* | *M* | *SD* | *M* | *SD* |
| Age (years) | 76.1* | 6.9 | 74* | 6.6 | 74.9 | 6.8 |
| Education (years) | 16.7 | 2.8 | 16.1 | 3 | 16.4 | 2.9 |
|  | *N* | % | *N* | % | *N* | % |
| Race |  |  |  |  |  |  |
| White | 86 | 96.6 | 108 | 91.5 | 194 | 93.7 |
| African American | 2 | 2.3 | 8 | 6.8 | 10 | 4.8 |
| Asian | 1 | 1.1 | 2 | 1.7 | 3 | 1.5 |
| CDR = 0 | 63 | 70.8 | 98 | 83.1 | 161 | 77.78 |
| Actigraphy | *M* | *SD* | *M* | *SD* | *M* | *SD* |
| TST (minutes) | 409.7 | 68.3 | 419.8 | 60.4 | 415.5 | 64 |
| SE (%) | 85.4 | 7 | 86.6 | 6 | 86.1 | 6.4 |
| PSQI | 4.2 | 3.2 | 4.9 | 3.6 | 4.6 | 3.4 |
| Verbal Memory | 0.4 * | 1.3 | 0.9 * | 1.2 | 0.7 | 1.3 |
| Executive Function | 0.1 | 0.9 | 0.4 | 0.8 | 0.3 | 0.8 |
| Attention | 0.1 | 0.4 | 0.2 | 0.4 | 0.2 | 0.4 |

CDR = Clinical Dementia Rating. TST = total sleep time. PSQI = Pittsburgh Sleep Quality Index. SE = sleep efficiency. M = mean. SD = standard deviation.
\* Differences between males and females, $p < 0.05$

### Sleep and Cognitive Function

The GAMs revealed some significant correlations between sleep parameters and cognitive function in older adults, after accounting for age, education, and CDR score. The degree of linearity can be determined by the effective degrees of freedom (EDF) value. An EDF value greater than 1 indicates the presence of non-linear patterns. In older females, total sleep time (TST) showed a non-linear association with executive function (edf = 2.10, *p* = 0.02; Figure 1) and verbal memory (edf = 1.39, *p* = 0.03; Figure 2). In executive function, this relationship demonstrated stable executive function performance up to a TST of approximately 400 minutes, beyond which a decrease in executive function was observed. The pattern of the relationship varied in verbal memory in which the decline in performance was more consistent with a longer TST. Additionally, TST presented a negative linear relationship with attention (edf = 1.00, *p* = 0.07). This association indicated that increasing TST was correlated with lower performance, although the relationship did not reach statistical significance (Fig 3). Among older males, TST showed negative linear relationships with cognitive performance across the three domains: executive function (edf = 1.00; *p* = 0.17; Figure 4), verbal memory (edf = 1.00; *p* = 0.07; Figure 5), and attention (edf = 1.00; *p* = 0.22; Figure 6). However, these associations were not statistically significant.

**Fig 1.**
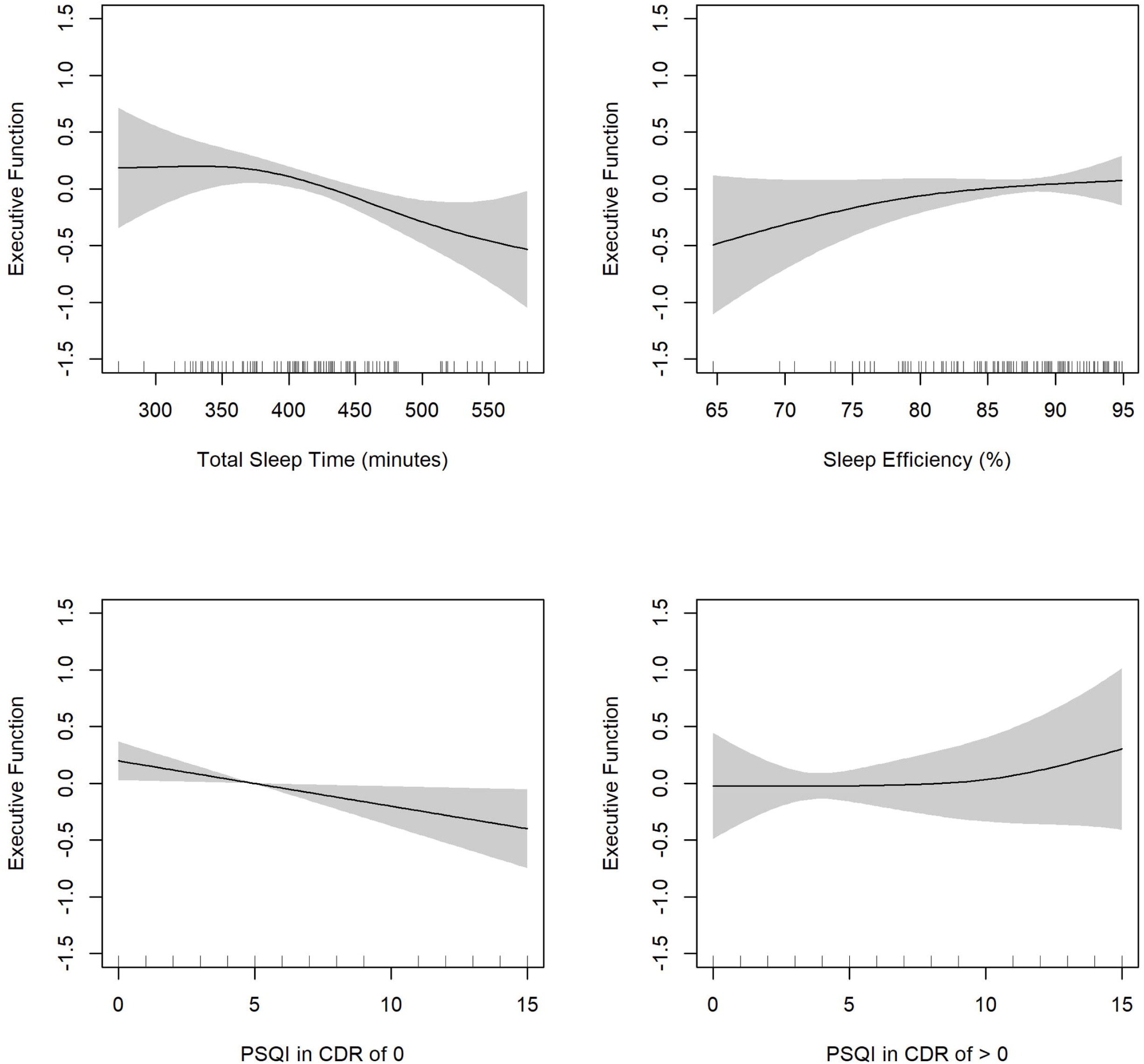
Partial effects of total sleep time, sleep efficiency, and Pittsburgh Sleep Quality Index (PSQI) on executive function in females.

**Fig 2.**
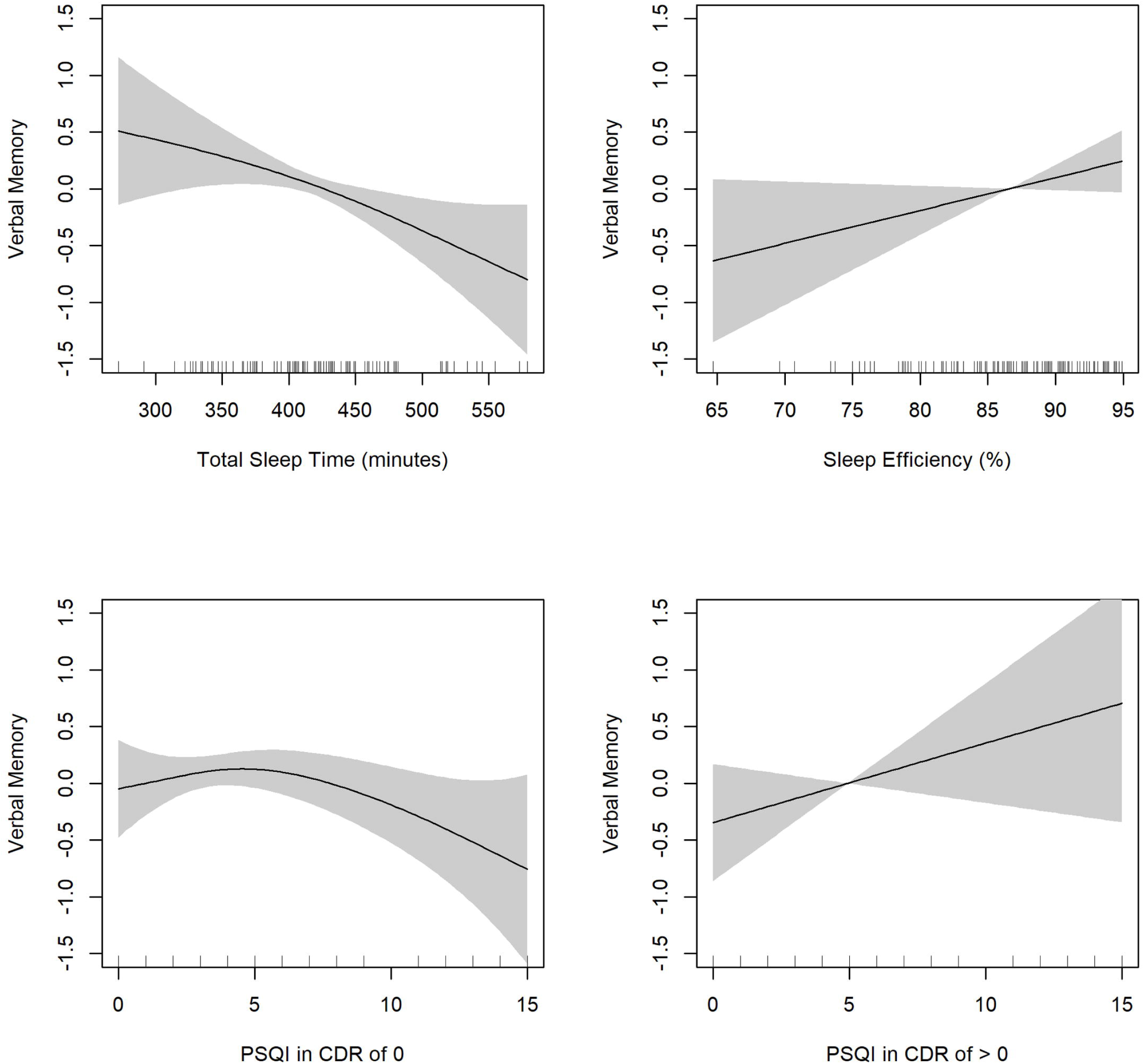
Partial effects of total sleep time, sleep efficiency, and Pittsburgh Sleep Quality Index (PSQI) on verbal memory in females.

**Fig 3.**
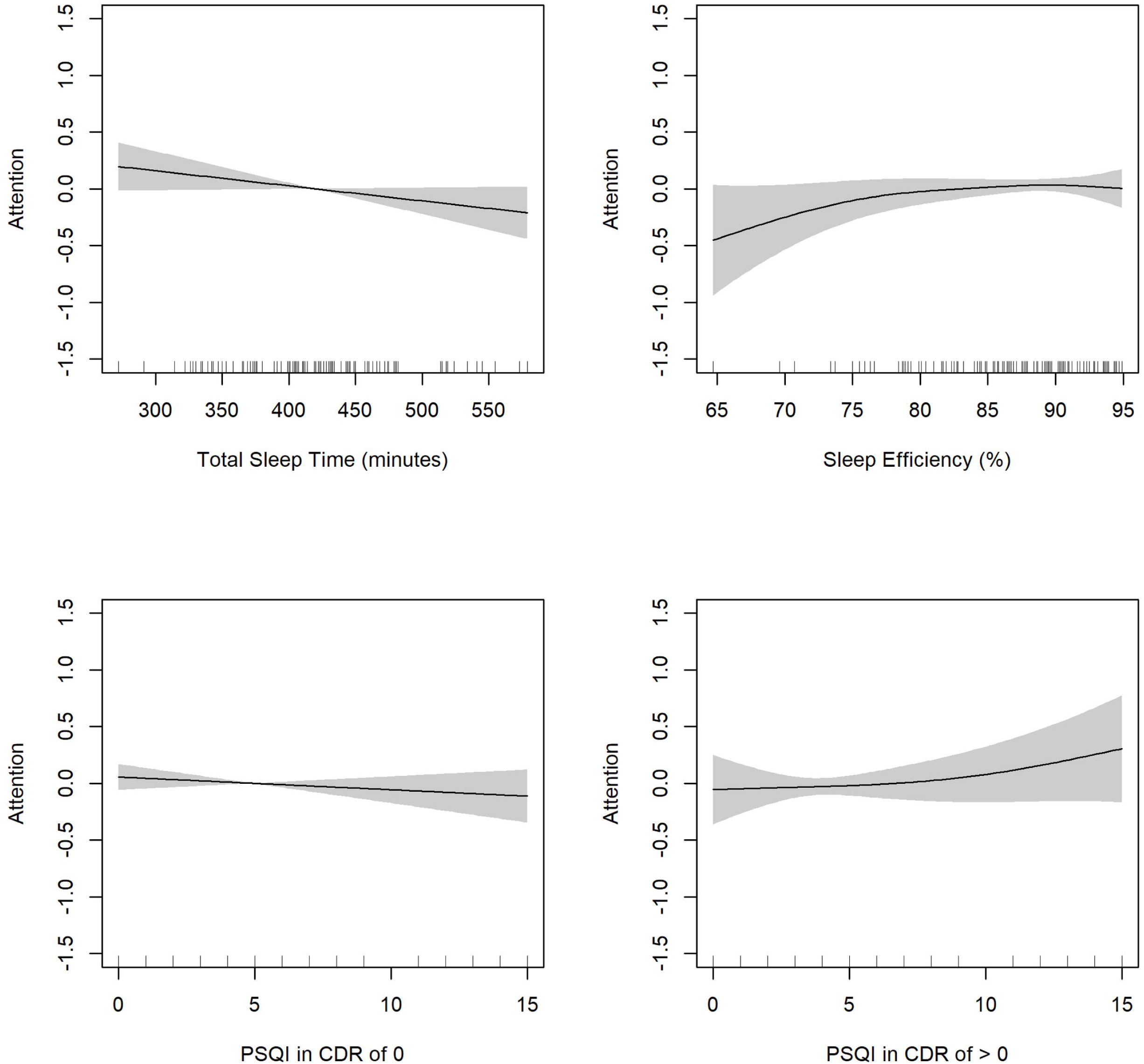
Partial effects of total sleep time, sleep efficiency, and Pittsburgh Sleep Quality Index (PSQI) on attention in females.

**Fig 4.**
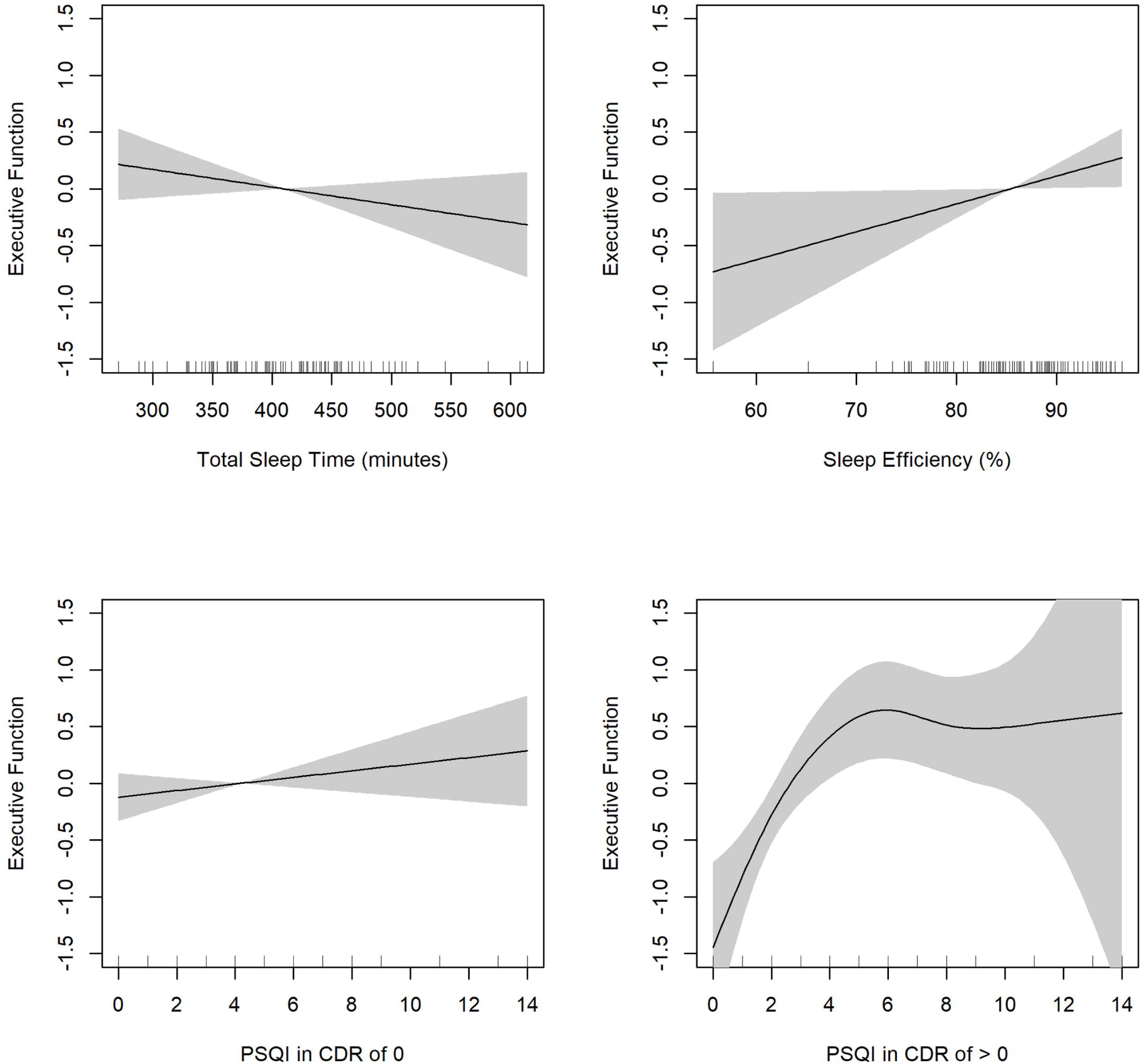
Partial effects of total sleep time, sleep efficiency, and Pittsburgh Sleep Quality Index (PSQI) on executive function in males.

**Fig 5.**
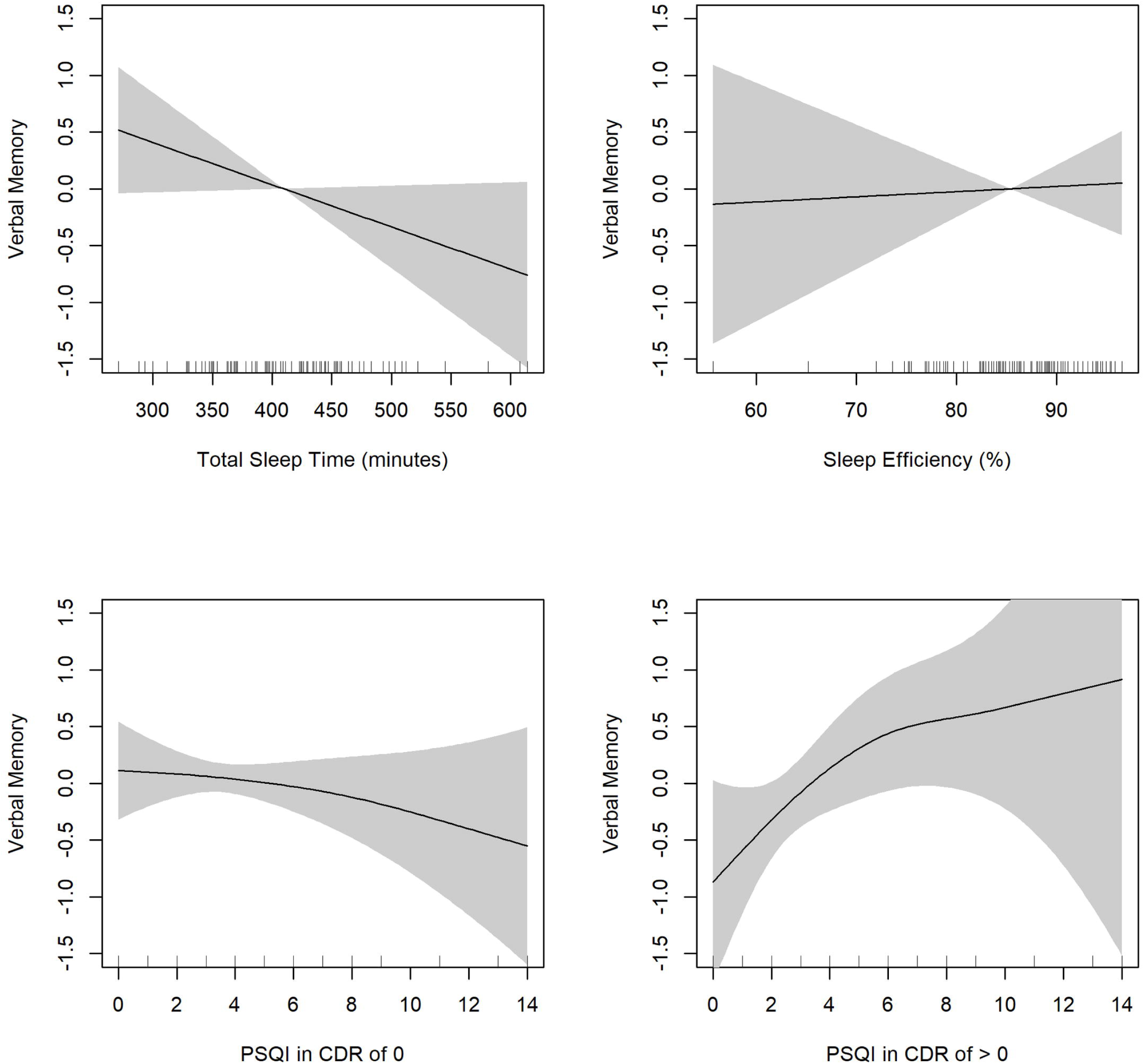
Partial effects of total sleep time, sleep efficiency, and Pittsburgh Sleep Quality Index (PSQI) on verbal memory in males.

**Fig 6.**
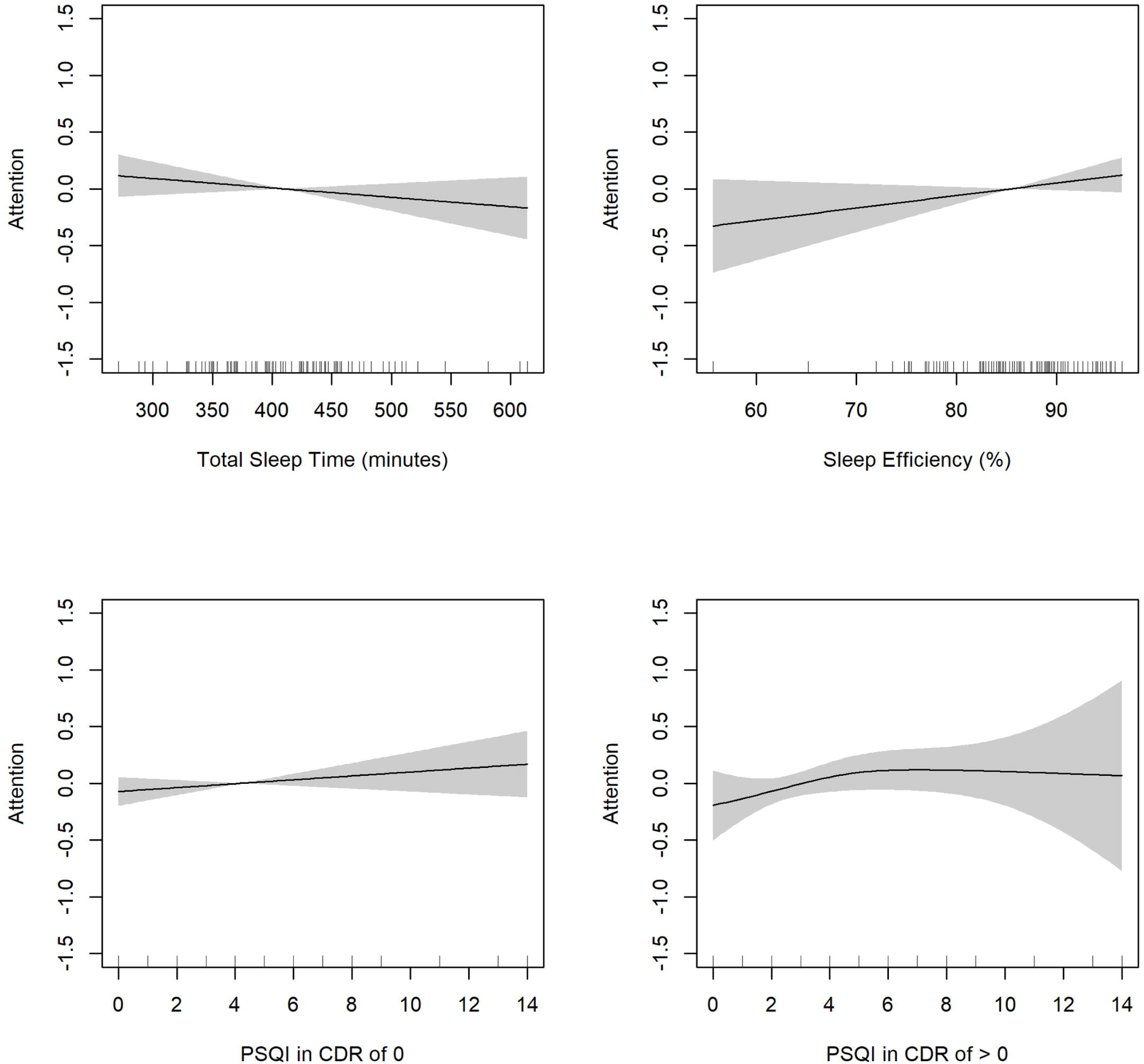
Partial effects of total sleep time, sleep efficiency, and Pittsburgh Sleep Quality Index (PSQI) on attention in males.

Sleep efficiency demonstrated non-linear associations with both executive function (edf = 1.57; *p* = 0.29) and attention (edf = 2.10; *p* = 0.29), while it was linearly related to verbal memory (edf =1.00; *p* =0.08) in older females (Figs 1-3). The overall pattern demonstrated that a higher SE was correlated with a higher performance. However, these associations did not reach statistical significance. Intriguingly, in older males, sleep efficiency positively correlated with executive function in a significant and linear manner (edf = 1.00, *p* = 0.04; Figure 4). The same pattern was observed between sleep efficiency and both verbal memory (edf = 1.00, *p* = 0.83) and attention (edf = 1.00, *p* = 0.12) in this group, although these relationships were not significant (Figs 5-6).

Total PSQI scores showed non-linear associations with verbal memory (edf = 1.97; *p* = 0.17; Figure 2) in older females with NC (CDR = 0). This relationship was marked by a decrease in verbal memory performance when the PSQI score exceeded 5. In addition, the PSQI had a linear negative relationship with executive function (edf = 1.00; *p* = 0.02; Figure 1) and attention (edf =1.00; *p* = 0.33; Figure 3). However, the relationship was only significant with executive function indicating poorer self-reported sleep quality, marked by a higher PSQI score, was associated with lower performance. In males with NC, the PSQI had a linear positive relationship with executive function (edf = 1.00; *p* = 0.26; Figure 4) and attention (edf = 1.00; *p* = 0.25; Figure 6) and a non-linear negative pattern with verbal memory (edf = 1.30; *p* = 0.52; Figure 5). These relationships were not statistically significant.

Among females with MCI, a linear negative relationship emerged between the PSQI and verbal memory (edf = 1.00; *p* = 0.18; Figure 5), and the PSQI exhibited non-linear positive relationships with executive function (edf = 1.46; *p* = 0.66; Figure 1) and attention (edf = 1.42; *p* = 0.41; Figure 3). Non-linear positive relationships were also observed in the PSQI with executive function (edf = 2.83, *p* = 0.001; Figure 4), verbal memory (edf = 1.58, *p* = 0.11; Figure 5), and attention (edf = 1.63; *p* = 0.37; Figure 6) in males with MCI, and these associations were only significant in executive function in males.

### Exploratory Results

We did not find significant correlations between objective and self-reported sleep parameters (TST and SE) in males and females with NC or MCI (Table 3). Moreover, we found that the original model (Model 4) showed the lowest AIC values for both males (179.38) and females (214.75) as shown in Table 4, suggesting that this model best explains the relationship between sleep and executive function. For verbal memory, the actigraphy-based model (Model 1) had the lowest AIC values for both males (282.03) and females (327.59), indicating that actigraphy-measured sleep parameters are more predictive of verbal memory performance. For Attention, the PSQI-based model (Model 2) exhibited the lowest AIC values for males (84.80), while the actigraphy-based model (Model 1) showed the lowest AIC values for females (118.45).

**Table 2.** Generalized Additive Models of the Relationship Between Sleep Quality (Total Sleep Time, Sleep Efficiency, and PSQI) and Cognitive Performance (Executive Function, Verbal Memory, and Attention)

|  | **Executive Function** |  | **Verbal Memory** |  | **Attention** |  |
| --- | --- | --- | --- | --- | --- | --- |
|  | Females | Males | Females | Males | Females | Males |
| **Spline fit** | **EDF** | **EDF** | **EDF** | **EDF** | **EDF** | **EDF** |
| TST | 2.10* | 1.00 | 1.39* | 1.00 | 1.00 | 1.00 |
| SE | 1.57 | 1.00* | 1.00 | 1.00 | 2.10 | 1.00 |
| PSQI (CDR=0) | 1.00* | 1.00 | 1.97 | 1.30 | 1.00 | 1.00 |
| PSQI (CDR>0) | 1.46 | 2.83** | 1.00 | 1.58 | 1.42 | 1.63 |
| **Covariate** | **Estimate** | **Estimate** | **Estimate** | **Estimate** | **Estimate** | **Estimate** |
| Age | -0.03** | -0.03** | -0.06*** | -0.03 | -0.02** | -0.02 |
| Education | 0.04* | 0.02 | 0.08** | 0.001 | 0.02 | 0.02 |
| CDR >0 | -0.94*** | -1.05*** | -1.18*** | -1.59*** | -0.32** | -0.44 |

Dependent variables: executive function, verbal memory, and attention. TST = total sleep time. SE = sleep efficiency. CDR = Clinical Dementia Rating. EDF = effective degrees of freedom (quantification of model's flexibility).
\* $p < 0.05$ , \*\* $p < 0.01$ , \*\*\* $p < 0.001$

**Table 3.** Spearman’s Correlations to Explore the Relationships Between TST and SE measured by Actigraphy and PSQI.

|  | Male |  | Female |  |
| --- | --- | --- | --- | --- |
|  | NC | MCI | NC | MCI |
| TST | -0.21 | -0.26 | -0.16 | 0.23 |
| SE | -0.20 | 0.22 | 0.04 | 0.24 |

TST = total sleep time. SE = sleep efficiency.
NC = normal cognition. MCI = mild cognitive impairment.

**Table 4.** Comparison of GAM Models Based on AIC Values for Different Sleep Measures.

|  | **Executive Function** |  | **Verbal Memory** |  | **Attention** |  |
| --- | --- | --- | --- | --- | --- | --- |
|  | Male | Female | Male | Female | Male | Female |
| Model 1 (Actigraphy) | 193.22 | 215.30 | 282.03 | 327.61 | 85.53 | 118.45 |
| Model 2 (PSQI) | 188.25 | 221.29 | 281.14 | 333.09 | 84.80 | 123.18 |
| Model 3 (Mix) | 189.24 | 218.85 | 281.52 | 331.97 | 86.33 | 123.06 |
| Model 4 (Original) | 179.38 | 214.75 | 281.84 | 327.59 | 86.98 | 120.50 |

## Discussion

Our study examined whether objective and self-reported sleep quality, as measured by actigraphy and a self-reported questionnaire, were associated with cognitive performance, specifically verbal memory, executive function, and attention, within an older adult clinical population. Concurrently, we investigated potential sex-related disparities in this association. We did not find significant differences in total sleep time, sleep efficiency, and self-reported sleep symptoms between the two sexes. Further research is needed to determine what constitutes optimal sleep for older females and males.

Based on our analysis, we found that total sleep time was a significant predictor, among older females but not older males, of executive function and memory, while attention was not associated in both sexes. The presence of both non-linear and linear relationships across various cognitive areas suggests that effects were not uniform but rather specific to distinct cognitive domains. This implies that each aspect of cognition may have its own threshold and optimal conditions for sleep duration and quality. To elucidate the sources of this heterogeneity, more research is required to unravel the complexities of how different cognitive functions are differentially affected by sleep and vice-versa. Meanwhile, low sleep efficiency was associated with lower executive function in older males. These findings suggest that sleep duration and sleep efficiency may have different effects on cognitive function between older females and males. Additionally, the results could indicate that sleep could have more pronounced effects on higher-order cognitive processes, such as memory and executive function ^20^ and less with attention that is considered a more basic cognitive process. Prior studies reported that acute sleep loss impacts different types of attention from the one focused on in our study, such as sustained attention, processing speed, and vigilance ^47,48^. It is noteworthy that our study concentrated on attention more closely aligned with working memory and short-term attention span, which could contribute to different findings from previous research.

We noted that those with MCI reported that more sleep disturbances tended to exhibit better cognitive performance, specifically executive function in males. This observation raises certain considerations. Existing research has proposed that those with cognitive impairment might have diminished self-awareness (i.e., anosognosia) ^45^, potentially leading to inaccuracies in their self-reports. This cautions against relying solely on self-report data when assessing individuals with cognitive impairment and highlights the importance of obtaining collateral information from caregivers. Another plausible explanation could be that sleep may no longer play as significant a role in cognition because these individuals may have already surpassed a critical threshold in their cognitive decline.

In terms of sex differences in perceiving sleep quality, previous research suggests that females generally have a more accurate perception ^26,27^. Our results suggested a trend among females with NC, where poorer self-reported sleep quality was associated with lower cognitive performance, while males with NC demonstrated more sporadic patterns between self-reported sleep quality and cognition. However, when comparing the total sleep time and sleep efficiency measured by actigraphy with self-reported data from our sample, we did not find significant correlations as indicated by the previous studies. This discrepancy could be because most of our sample scored in the "better" range on the sleep duration and efficiency subscales of the PSQI. To better understand the relationship between objective and self-reported sleep measures, future research should involve a larger and more diverse sample, including individuals with varying sleep quality.

One main difference between our study and most studies that investigate sleep and cognition is that prior research typically induces sleep loss to investigate the effects of acute or chronic sleep loss on cognitive performance, resulting in a significant impact on memory and executive function ^30,31,49^, as opposed to real-world observations in our study. Furthermore, the majority of our participants exhibited ideal sleep efficiency levels (around 85%). Studies that include those with lower sleep efficiency may yield different results and provide insights into the cognitive sequelae of clinically impaired sleep.

Our study has several strengths. First, we used a comprehensive neuropsychological battery to gauge participants’ cognitive performance, opting for a more in-depth assessment rather than relying on a simple test battery (e.g., the Mini-Mental State Examination), which is less sensitive to subtle cognitive impairment. Second, we included composite scores of cognitive domains generated through confirmatory factor analysis to represent each cognitive domain using multiple tests. This approach allows for a better representation of each cognitive domain. Third, we employed generalized additive models to capture non-linear relationships between sleep quality and cognitive performance without imposing symmetrical curves. Moreover, GAMs have the built-in capability to determine the appropriate level of smoothness for each predictor, ensuring an optimal fit. Fourth, we objectively measured total sleep time and sleep efficiency over multiple nights allowing for a more representative result of typical sleep and compared it to self-reported measures to evaluate distinctions between the two common types of measures.

Several limitations should also be acknowledged. First, the present study is limited by the characteristics of the sample. The participants were primarily White, highly educated, relatively healthy, and motivated to engage in research, leading to limited generalizability of the findings. Most of the participants in this study had relatively healthy sleep patterns. Second, we did not include daytime napping in the total sleep time because previous research suggests that daytime and nighttime sleep have different impacts on cognitive processes ^50^. Therefore, excluding daytime naps helped to isolate the specific effects of nighttime sleep, ensuring a clearer understanding of how it relates to cognitive performance. Third, we acknowledged the potential challenges of using the PSQI in individuals with MCI, particularly regarding their ability to recall sleep details over the past 30 days. Despite these concerns, we found that the instrument has been extensively used in research involving MCI populations as reported in a literature review ^51^, which supports our decision to include participants with normal and mildly impaired cognition in our analysis, allowing for a more comprehensive exploration of the relationship between sleep quality and cognitive function across a spectrum of cognitive statuses. Moreover, the potential influence of cognitive impairment on self-reported sleep quality was mitigated by incorporating the clinical dementia rating (CDR) into the analysis as a covariate. We also stratified the relationship between PSQI and cognitive outcomes by CDR levels.

Our study has yielded foundational evidence of the association between sleep and cognitive health in older females and males, with effects across distinct cognitive domains. Our findings suggest that studies focused on determining the optimal sleep characteristics required to maintain cognitive health in individuals of different sexes are warranted. Future research should also explore the long-term effects of midlife hormonal changes on sleep and cognitive health in aging populations, using longitudinal analysis, since this period is notable for hormonal shifts. Midlife may serve as an optimal window for intervention, as cognitive decline resulting from sleep deprivation tends to be less severe at this stage. Such studies could also provide critical insights into the temporal dynamics of sleep and cognitive decline, identifying potential periods of vulnerability and opportunities for preventive interventions. Moreover, future research should incorporate information regarding stages of sleep measured by polysomnography (PSG) to provide a more detailed understanding of how sleep architecture contributes to cognitive function and whether these effects vary by sex. The implications of future findings have the potential to shape public health policies and strategies for aging. Healthcare professionals should consider recommending regular sleep assessments as part of a comprehensive check-up to promote cognitive well-being in aging populations.

## Data Availability

The dataset presented in this article is not readily available because dataset requests must be made directly to the KU-ADRC. Those interested in accessing the dataset should be directed to the following website where they can complete a data request form: https://www.kumc.edu/research/alzheimers-disease-research-center/research/resources-for-researchers-and-principal-investigators.html.

